# Clinician knowledge and self-efficacy in snakebite management: A cross-sectional assessment in Northern Uganda

**DOI:** 10.64898/2026.06.15.26355658

**Authors:** Dickens Kamugisha, Phillip Musoke, Nelson Ssewante, David Musoke, Felix Bongomin

**Author notes:** **Correspondence:** (DK); (PM); (FB).

## Abstract

**Background:** Snakebite envenomation (SBE) is a major public health crisis in rural Uganda, yet it remains a neglected tropical disease. Effective management is often compromised by systemic barriers and a lack of clinician training. This study assessed clinician self-efficacy and objective knowledge regarding SBE management in Northern Uganda.

**Methods:** A descriptive, cross-sectional study was conducted between February and July 2025 among 379 healthcare workers in Gulu, Omoro, and Pader districts. A validated questionnaire was used to collect data on socio-demographics, self-reported efficacy (scale 1–10), and objective knowledge. Knowledge scores ≥70% were categorized as adequate. Multivariable logistic regression identified independent predictors of adequate knowledge, and Spearman’s correlation (ρ) assessed the relationship between knowledge and self-efficacy.

**Results:** The participants had a mean age of 35.6 years (SD ±7.3), were predominantly female (56.5%, 214/379), and most (83.6%, 317/379) practiced at Health Centre III level facilities. While 53.8% (204/379) reported prior training, 48.3% (183/379) of these had not received an update in over 10 years. Adequate knowledge was demonstrated by 51.5% (195/379) of participants. In the multivariable analysis, practicing in Omoro (adjusted odds ratio [aOR]: 0.3, 95% CI: 0.1–0.6, p < 0.001) or Pader (aOR: 0.2, 95% CI: 0.1–0.4, p < 0.001) was associated with lower odds of adequate knowledge compared to Gulu district. Prior training significantly increased the odds of adequate knowledge (aOR: 2.3, 95% CI: 1.3–4.2, p = 0.006). A moderate positive correlation was observed between self-efficacy and objective knowledge (Spearman’s ρ = 0.33, p < 0.0001).

**Conclusion:** Approximately half of the frontline healthcare workers in Northern Uganda lack adequate knowledge on SBE management, with significant geographic differences and outdated training. The gap between clinician self-efficacy and objective knowledge poses a risk to patient safety. Regular, mandatory refresher training and targeted educational outreach to remote districts are required to reduce SBE-related morbidity and mortality.

## Introduction

Snakebite envenomation (SBE) is a serious but often overlooked worldwide public health issue that requires immediate improvements in prevention, clinical care, and rehabilitation (1). Up to 2.7 million people experience envenomation each year, with a shocking death rate of 81,000 and 138,000 (2). The great majority of these instances take place in poor rural communities where people frequently come into contact with poisonous snakes due to subsistence activities (2). An estimated 7,000 to 32,000 people die each year in sub-Saharan Africa (SSA) (3); however, as over 70% of cases go unreported, these numbers probably only reflect a small portion of the actual burden (4,5). In Uganda, where a high percentage of the population resides in rural areas and relies on agriculture, the risk of SBE is particularly significant.

An untreated snakebite has a catastrophic clinical course. Victims frequently experience multi-system consequences in addition to imminent death, such as rhabdomyolysis, neurotoxicity, acute renal injury, respiratory failure, and local tissue damage (6–9). The long-term consequences are equally devastating, including severe psychological distress, blindness, maternal-fetal loss, and physical disabilities such as contractures and amputations (2,10). Effective care is hindered by a number of systemic barriers, even though prompt delivery of the right antivenom is the gold standard for management (6,11). Inadequate cold-chain storage facilities, variable antivenom supply chains, and product efficacy concerns sometimes hinder treatment in resource-constrained settings (11,12).

The effectiveness of the medical response is greatly dependent on the competence of frontline clinicians, who serve as the primary point of contact for victims (13). Despite their key role, evidence suggests a deficiency of specialized knowledge and practical skills in SBE management among healthcare workers (14). Data about healthcare workers’ knowledge and skills in SBE is still scarce in SSA. However, a recent study indicated that only 12% of healthcare workers had received formal training in snakebite management (15). This knowledge gap is reflected in neighboring SSA nations, where persistent misconceptions and insufficient clinical training continue to compromise patient outcomes (15–17).

Addressing this challenge requires a two-pronged strategy that targets both institutional resources and human capital. There is a need to move beyond anecdotal reports and systematically assess the actual availability and storage integrity of antivenoms in rural health centers. Furthermore, understanding the relationship between a clinician’s self-reported efficacy and their objective clinical knowledge is vital for developing effective educational interventions.

This study addressed these gaps by assessing the current state of clinician readiness for SBE management in Northern Uganda. Specifically, it evaluated frontline healthcare personnel’s self-reported efficacy levels and compared them with objective clinical knowledge of snakebite management.

## Methods

### Ethical Statement

The study protocol was reviewed and approved by the Mulago Hospital Research and Ethics Committee (Reference Number: MHREC 2856). The study was conducted in strict accordance with the principles of the Declaration of Helsinki.

### Study Design

A descriptive, health facility-based cross-sectional survey was employed to assess the readiness of frontline clinicians in managing snakebite envenomation (SBE). This manuscript was prepared in accordance with the STROBE (Strengthening the Reporting of Observational Studies in Epidemiology) (18) statement for cross-sectional studies **(supplementary material 1)**.

### Study Setting

The research was situated in Gulu, Omoro, and Pader districts of Northern Uganda. This region is characterized by a unique ecological profile; unlike the rest of the country which experiences bimodal rainfall, this area follows a unimodal tropical climate with a single rainy season extending from March to October (19). Ambient temperatures fluctuate between 15°C and 30°C, providing an ideal environment for medically important snake species, including the black mamba (*Dendroaspis polylepis*), puff adder (*Bitis arietans*), and various cobras (*Naja spp*.) (20). The socio-economic landscape is predominantly agricultural and rural. The combination of dense vegetation during the rains and the prevalence of subsistence farming significantly elevates the risk of human-snake encounters. Consequently, these districts are recognized as SBE-endemic zones where healthcare facilities serve as critical lifelines for bite victims (21).

### Study Population

The target population included clinical officers, medical officers, and specialist physicians active in patient care during the study period in the three districts. To ensure a representative sample of rural healthcare facilities, we recruited participants from Health Centre IIIs, Health Centre IVs, and district hospitals.

### Sampling and Eligibility

A sample size of 422 clinicians was calculated to provide 80% power at a 95% confidence level, assuming a 50% baseline self-efficacy and a 5% type I error margin. We included clinicians over the age of 18 who provided direct patient care and gave informed consent. To maintain data integrity, we excluded visiting clinicians, medical students, and administrative staff not involved in active clinical management.

### Data Collection Procedures

Data were collected between February 2025 and July 2025 using an electronic questionnaire developed in KoboToolbox. The items used to assess knowledge of snakebite management were developed from the tool used to assess Healthcare workers’ knowledge of the management and treatment of snakebite cases in rural Malawi (22). Trained research assistants administered the survey face-to-face. The instrument comprised three primary sections (supplementary material 2):

### Socio-demographics

Capturing age, sex, education level, facility level, and years of professional experience.

### Self-Efficacy

Clinicians rated their confidence in managing SBE on a scale of 1 (“not confident at all”) to 10 (“totally confident”).

### Objective Knowledge

Questions were adapted from validated tools used in similar sub-Saharan African contexts (22). This section included questions on knowledge of snakebite management.

### Data Analysis and Management

Data analysis was performed using STATA (StataCorp, College Station, TX, USA). Upon completion of data collection, a final dataset (supplementary material 3) was downloaded and imported into STATA for cleaning and formal analysis. A composite knowledge score was generated by summing responses to the knowledge items in the original questionnaire. The total score was converted into a percent score and then dichotomized into a binary outcome variable categorized as “adequate” and “inadequate” knowledge, using 70% as the cutoff. Descriptive statistics were used to summarize participant characteristics. Continuous variables were summarized using means and standard deviations (SD) where normally distributed, while skewed variables were summarized using medians and interquartile ranges (IQR). Categorical variables were summarized using frequencies and percentages. Cross-tabulations were performed, and the Chi-square test (or Fisher’s exact test where appropriate) was used to compare proportions between groups. Variables with a p<0.20 at the bivariate level were considered for inclusion in the multivariable logistic regression model. A multivariable logistic regression model was fitted to identify independent factors associated with adequate knowledge of snakebite management. Adjusted odds ratios (aORs) with 95% confidence intervals (CIs) were reported as measures of association. For self-efficacy, a composite score was calculated by taking the average of the three items. The final score was not normally distributed and the median and interquartile range (IQR) were used. The relationship between self-efficacy and knowledge scores was assessed using Spearman’s rank correlation coefficient (ρ), as both variables were either ordinal or not normally distributed. Statistical significance was set at p < 0.05.

## Results

### Characteristics of study participants

A total of 379 healthcare workers participated in the study. The mean age of participants was 35.6 years (SD ±7.3), with the majority aged 26–35 years (42.0%), followed by 36–45 years (38.0%). Only 9.8% were aged 18–25 years, while 10.3% were older than 45 years. More than half of the participants were female (56.5%), and the majority held a diploma-level qualification (61.5%), followed by certificate holders (30.9%) and a small proportion with bachelor’s degrees (7.7%).

Most participants were working in Health Centre III facilities (83.6%), with fewer in Health Centre IV (13.5%) and hospitals (2.9%). The distribution across districts was relatively even, with approximately one-third of participants from Gulu (33.5%), Omoro (33.3%), and Pader (33.3%).

The median duration of professional experience was 10 years (IQR: 7–13), with the largest proportion having 6–10 years (43.8%) or more than 10 years of experience (38.0%). Slightly more than half (55.4%) had never administered SAV, while 53.8% reported having received prior training in snakebite management. Among those trained, the median duration since last training was 3 years (IQR: 2–6), although nearly half (48.3%) had been trained more than 10 years prior (Table 1).

**Table 1:** Characteristics of healthcare workers in Omoro, Pader, and Gulu Districts in Northern Uganda.

| Variable | n (%) |
| --- | --- |
| **Age, mean (SD)** | **35.6(7.3)** |
| 18-25 years | 37(9.8) |
| 26-35 years | 159(42.0) |
| 36-45 years | 144(38.0) |
| >45 years | 39(10.3) |
| **Sex** |  |
| Female | 214(56.5) |
| Male | 165(43.5) |
| **Level of education** |  |
| Bachelors | 29(7.7) |
| Certificate | 117(30.9) |
| Diploma | 233(61.5) |
| **Facility level** |  |
| Health centre III | 317(83.6) |
| Health centre IV | 51(13.5) |
| Hospital | 11(2.9) |
| **District where facility is located** |  |
| Gulu | 127(33.5) |
| Omoro | 126(33.3) |
| Pader | 126(33.3) |
| **Professional experience, Median (IQR)** | **10 (7,13)** |
| One year | 18(4.8) |
| 2-5 years | 51(13.5) |
| 6-10 years | 166(43.8) |
| >10 years | 144(38.0) |
| **Ever administered SAV** |  |
| No | 210(55.4) |
| Yes | 169(44.6) |
| **Ever been trained in snake bite management** |  |
| No | 175(46.2) |
| Yes | 204(53.8) |
| **Duration since last training, Median (IQR)** | **3 (2,6)** |
| One year | 31(8.2) |
| 2-5 years | 121(31.9) |
| 6-10 years | 44(11.6) |
| >10 years | 183(48.3) |

### Knowledge of snakebite management and antivenom use

A total of 195 (51.5%) participants had adequate knowledge. Overall, knowledge of snakebite management varied significantly across participant characteristics (Table 2). Age was significantly associated with knowledge levels (p < 0.001), with higher proportions of adequate knowledge observed among older participants. For example, 71.8% of those aged >45 years had adequate knowledge compared to 37.8% among those aged 18–25 years. Sex was also associated with knowledge (p = 0.012), with a higher proportion of males (58.8%) demonstrating adequate knowledge compared to females (45.8%). Education level showed a strong association (p < 0.001), with participants holding bachelor’s degrees (62.1%) and diplomas (59.7%) more likely to have adequate knowledge compared to certificate holders (32.5%). Geographical differences were notable (p < 0.001). Participants from Gulu had the highest proportion of adequate knowledge (71.7%), followed by Omoro (50.8%), while Pader had the lowest (31.7%). Professional experience was significantly associated with knowledge (p = 0.004), with increasing experience corresponding to higher knowledge levels. Similarly, participants who had ever administered SAV (60.4% vs 44.3%, p = 0.002) and those who had received prior training (61.3% vs 40.0%, p < 0.001) were more likely to demonstrate adequate knowledge. Recency of training was also important (p = 0.002), with higher knowledge levels observed among those trained more recently.

**Table 2:** Bivariate analysis of participant characteristics and knowledge about snakebite antivenom among healthcare workers in Omoro, Pader and Gulu Districts in Northern Uganda.

| Variable | Adequate<br>n (%) | Inadequate<br>n (%) | p-values |
| --- | --- | --- | --- |
| **Age** |  |  | **<0.001** |
| 18-25 years | 14(37.8) | 23(62.2) |  |
| 26-35 years | 61(38.4) | 98(61.6) |  |
| 36-45 years | 92(63.9) | 52(36.1) |  |
| >45 years | 28(71.8) | 11(28.2) |  |
| **Sex** |  |  | **0.012** |
| Female | 98(45.8) | 116(54.2) |  |
| Male | 97(58.8) | 68(41.2) |  |
| **Level of education** |  |  | **<0.001** |
| Bachelors | 18(62.1) | 11(37.9) |  |
| Certificate | 38(32.5) | 79(67.5) |  |
| Diploma | 139(59.7) | 94(40.3) |  |
| **Facility level** |  |  | **0.407** |
| Health centre III | 161(50.8) | 156(49.2) |  |
| Health centre IV | 26(51.0) | 25(49.0) |  |
| Hospital | 8(72.7) | 3(27.3) |  |
| **District where facility is located** |  |  | **<0.001** |
| Gulu | 91(71.7) | 36(28.3) |  |
| Omoro | 64(50.8) | 62(49.2) |  |
| Pader | 40(31.7) | 86(68.3) |  |
| **Professional experience** |  |  | **0.004** |
| One year | 4(22.2) | 14(77.8) |  |
| 2-5 years | 21(41.2) | 30(58.8) |  |
| 6-10 years | 83(50.0) | 83(50.0) |  |
| >10 years | 87(60.4) | 57(39.6) |  |
| **Ever administered SAV** |  |  | **0.002** |
| No | 93(44.3) | 117(55.7) |  |
| Yes | 102(60.4) | 67(39.6) |  |
| **Ever been trained in snake bite management** |  |  | **<0.001** |
| No | 70(40.0) | 105(60.0) |  |
| Yes | 125(61.3) | 79(38.7) |  |
| **Duration since last training** |  |  | **0.002** |
| One year | 21(67.7) | 10(32.3) |  |
| 2-5 years | 73(60.3) | 48(39.7) |  |
| 6-10 years | 25(56.8) | 19(43.2) |  |
| >10 years | 76(41.5) | 107(58.5) |  |

### Multivariable analysis

After adjusting for potential confounders, the district of practice and training in snakebite management remained significantly associated with knowledge.

Compared to participants from Gulu, those from Omoro (aOR = 0.3; 95% CI: 0.1–0.6; p < 0.001), and Pader (aOR = 0.2; 95% CI: 0.1–0.4; p < 0.001) had significantly lower odds of adequate knowledge. Participants who had received prior training in snakebite management were more than twice as likely to have adequate knowledge compared to those without training (aOR = 2.3; 95% CI: 1.3–4.2; p = 0.006). Other variables such as age, sex, education level, professional experience, and prior administration of SAV were not statistically significant after adjustment (Table 3).

**Table 3:** Logistic regression of the factors associated with knowledge about snakebite antivenom among healthcare workers in Omoro, Pader and Gulu Districts in Northern Uganda.

| Variable | cOR (95% CI) | p-value | aOR (95% CI) | p-value |
| --- | --- | --- | --- | --- |
| **Age** |  |  |  |  |
| 18-25 years | 1.0 |  |  |  |
| 26-35 years | 1.0(0.5,2.1) | 0.953 | 0.8(0.3,2.3) | 0.631 |
| 36-45 years | 2.9(1.4,6.1) | 0.005 | 1.6(0.5,5.4) | 0.450 |
| >45 years | 4.2(1.6,11.0) | 0.004 | 4.0(0.8,19.7) | 0.087 |
| **Sex** |  |  |  |  |
| Female | 1.0 |  |  |  |
| Male | 1.7(1.1,2.5) | 0.012 | 1.5(0.9,2.4) | 0.140 |
| **Level of education** |  |  |  |  |
| Certificate | 1.0 |  |  |  |
| Diploma | 3.1(1.9,4.9) | <0.001 | 1.1(0.6,2.2) | 0.681 |
| Bachelors | 3.4(1.5,7.9) | 0.004 | 0.6(0.2,2) | 0.387 |
| **District where facility is located** |  |  |  |  |
| Gulu | 1.0 |  |  |  |
| Omoro | 0.4(0.2,0.7) | 0.001 | 0.3(0.1,0.6) | **<0.001** |
| Pader | 0.2(0.1,0.3) | <0.001 | 0.2(0.1,0.4) | **<0.001** |
| **Professional experience** |  |  |  |  |
| One year | 1.0 |  |  |  |
| 2-5 years | 2.4(0.7,8.5) | 0.158 | 2.5(0.6,9.8) | 0.182 |
| 6-10 years | 3.5(1.1,11.1) | 0.033 | 1.4(0.3,7) | 0.647 |
| >10 years | 5.3(1.7,17.0) | 0.005 | 1.2(0.2,6.5) | 0.822 |
| **Ever administered SAV** |  |  |  |  |
| No |  |  |  |  |
| Yes | 1.9(1.3,2.9) | 0.002 | 0.9(0.5,1.8) | 0.835 |
| **Ever been trained in snake bite management** |  |  |  |  |
| No | 1.0 |  |  |  |
| Yes | 2.4(1.6,3.6) | <0.001 | 2.3(1.3,4.2) | **0.006** |

### Self-reported self-efficacy for diagnosis and management of snakebites

The median self-efficacy score among participants was 10 (IQR: 1–10). There was a moderate positive correlation between self-efficacy and knowledge scores (Spearman’s ρ = 0.3313, p < 0.0001), suggesting that participants with higher knowledge levels were more likely to report greater confidence in managing snakebite cases.

## Discussion

This study offers an important assessment of clinician readiness to manage SBE in Northern Uganda, revealing a significant gap between professional self-efficacy and objective clinical competence. Our findings show that while over half of the participants reported high self-efficacy, only just over half (fifty-one percent) possessed adequate knowledge of SBE management. Importantly, the multivariable analysis identified geographical location and prior training as the only independent predictors of knowledge, highlighting systemic inequities in professional development across the region.

Only 51.5% of clinicians demonstrated adequate knowledge, which is consistent with similar studies conducted in sub-Saharan Africa, where SBE remains a neglected topic. For example, research conducted in northern Nigeria and Kenya revealed similar knowledge gaps among frontline workers (15,17). However, our cohort’s “training paradox”—nearly half of the previously trained health workers having not received an update in more than ten years—suggests that one-off training is inadequate. The significant association between training and knowledge aligns with findings from Cameroon, which emphasized that targeted educational interventions are the most effective means of improving SBE outcomes (16). Our findings highlight that without regular refreshers, clinical protocols for neglected tropical diseases quickly become obsolete. Clinicians in Pader had 80% and Omoro had 70% lower odds of adequate knowledge compared to Gulu. This may suggest a “knowledge drain” away from urban centers. Gulu’s proximity to major referral hospitals and academic institutions like Gulu University may be beneficial, yet the more rural districts appear neglected. This suggests that in Northern Uganda, institutional support may be concentrated in specific hubs, leaving rural clinicians at Health Centre III levels who see the majority of victims unprepared.

The moderate positive correlation between self-efficacy and objective knowledge is perhaps the most clinically concerning finding. A significant portion of our participants reported maximum confidence (median score 10) despite failing to meet the 70% knowledge threshold. This “confidence-competence gap” suggests that medical professionals may be using out-of-date or inaccurate assumptions to make important medical decisions like wound debridement or antivenom dosage. This behaviour is a significant risk factor for medical mistakes and preventable mortality and has been observed in various emergency care settings (23).

These findings have direct implications for public health policy in Uganda. The Ministry of Health should consider SBE management as a mandatory component of Continuing Medical Education (CME), specifically targeting rural districts like Pader and Omoro. Furthermore, our results suggest that interventions should not just focus on the presence of antivenom, but on the human capital required to administer it. Methodologically, this study shows that objective testing is still the gold standard for determining competency; self-efficacy ratings by themselves are insufficient criteria for measuring clinician readiness.

Several limitations must be acknowledged. First, the study’s cross-sectional design makes it impossible to establish a causal relationship between knowledge and training. Second, although the 70% limit for “adequate knowledge” is a widely used academic standard, it might not adequately reflect the sophisticated abilities needed in complicated envenomation cases. Finally, our sample was slightly below the calculated target of 422, though 379 participants still provided sufficient power for robust multivariable analysis (response rate was 85%). We minimized potential bias by using face-to-face interviews to ensure participants did not consult external materials during the assessment.

This study shows a need for standardized and regular snakebite management training in Northern Uganda. Although training is a strong predictor of knowledge, its influence gradually diminishes. Reducing the incidence of snakebite mortality in rural communities requires addressing regional inequities and bridging the gap between clinician confidence and real ability.

## Conclusions

This study reveals that while healthcare workers in Northern Uganda possess a moderate level of confidence in managing snakebite envenomation, their objective clinical knowledge is inconsistent and is mostly reliant on how recently they received professional training. Clinicians in the most distant districts may be considerably less equipped to handle these medical situations than those in more urbanized areas. Furthermore, the observed gap between self-reported efficacy and actual knowledge indicates a risk of clinical overconfidence that could compromise patient safety. These findings highlight the necessity of moving beyond one-off training sessions toward a system of regular, standardized clinical updates. Improving snakebite outcomes and lowering preventable deaths in rural Uganda requires strengthening the human-resource capacity through focused, district-specific educational interventions. This study identifies a significant gap between professional confidence and actual competence in SBE management among healthcare workers in Northern Uganda. This disparity emphasizes the need for mandatory, regular refresher training and standardized clinical protocols to reduce avoidable mortality in rural communities.

## Data Availability

Pooled outcome data and analysis code are available from the corresponding author on reasonable request. Individual participant data from the included trials are subject to the respective trial data sharing agreements.

## Supporting information

Supplementary material 1. **STROBE Checklist for Cross-Sectional Studies**. von Elm E, Altman DG, Egger M, Pocock SJ, Gøtzsche PC, Vandenbroucke JP. The Strengthening the Reporting of Observational Studies in Epidemiology (STROBE) statement: guidelines for reporting observational studies. J Clin Epidemiol [Internet]. 2008 Apr;61(4):344–9. Available from: pmid:18313558, Doi: 10.1016/j.jclinepi.2007.11.008

Supplementary material 2. **Questionnaire**.

Supplementary material 3. **Detailed Questionnaire Responses**.

Supplementary material 4. **STATA output**

## Acknowledgement

We express our sincere gratitude to the District Health Offices of Gulu, Omoro, and Pader for their administrative support and for granting permission to conduct this study within their jurisdictions. We are deeply indebted to the management and staff of the participating health facilities, whose cooperation and commitment to improving snakebite care made this research possible. Special thanks are extended to our dedicated research assistants, Dr. Patrick Atiya and Ms. Fiona Lake, for their tireless efforts in data collection and ensuring the integrity of the field operations. We also thank the healthcare workers of Northern Uganda who generously shared their time and perspectives to inform this study. Lastly, we would like to express our gratitude for the small early career grant awarded by the Royal Society of Tropical Medicine & Hygiene (RSTMH) and the National Institute for Health and Care Research (NIHR) under Award Number (Grant code: nihr24033, Grant ID: 46296932). The funders had no role in study design, data collection and analysis, decision to publish, or preparation of the manuscript.

## Data Sharing Statement

The data used to support the results of the research are available from the corresponding author upon request.

## Author Contributions

All authors made substantial contributions to conception and design, acquisition of data, or analysis and interpretation of data, took part in drafting the article or revising it critically for important intellectual content, agreed to submit to the current journal, gave final approval to the version to be published, and agreed to be accountable for all aspects of the work.

